# Disparities and trends in global representation of human genetics conferences: a 26-year longitudinal study of ASHG and ESHG

**DOI:** 10.1101/2025.08.12.25333491

**Authors:** Hao Zheng, Ying Wang, Yosuke Tanigawa, Jue Sheng Ong, Stuart MacGregor, Liming Liang, Manolis Kellis, Xikun Han

## Abstract

Equity in human genetics research requires balanced participation not only from study participants from global populations but also from the researchers who drive the science. While disparities among research participants across ancestries and countries have been well studied, the representation and disparities of researchers themselves on the global stage remains poorly understood. Here, we analyzed over 100,000 abstracts presented at two leading annual conferences in the field, the American Society of Human Genetics (ASHG) and the European Society of Human Genetics (ESHG), from 1999 to 2024 to assess trends and geographic disparities. North America and Europe consistently dominated abstract contributions, whereas continents such as Africa, Oceania, and East Asia remained underrepresented, despite gradual increases in participation. The imbalance was even more pronounced in oral presentation: at ASHG, abstracts from North America were approximately 4 times more likely to be selected for talks than those from East Asia and 23 times more likely than those from South America; at ESHG, Europe’s advantage was 2 times and 9 times, respectively. Notably, Oceania had the highest relative success in oral presentation, with a ratio 5 times higher than East Asia and 29 times higher than South America in ASHG, and 8 times and 33 times higher, respectively, in ESHG. To explore potential drivers of these disparities, we examined 6 national level variables. The multivariable regression model indicated that GDP is the primary factor for abstract, while Nature Index Share is the main factor for oral presentation counts. Our findings highlight persistent global inequalities in representation of human geneticists at premier conferences. Greater international support and targeted initiatives are needed to promote more equitable worldwide involvement in human genetics.

## Introduction

The field of human genetics has long been characterized by global disparities in resources and research participation^1,2^. Key resources, such as global biobanks^3,4^, genome-wide association studies^5^ (GWAS), and polygenic risk scores^6^ (PRS), serve as critical roles in prediction medicine from a genome perspective^7^, yet access to and representation within these resources remain highly unbalanced across populations. Although the GWAS Catalog documented an increase in Asian and African ancestry representation between 2009 and 2016, European-ancestry datasets remained overwhelmingly predominant (81%), with Hispanic/Latin American ancestries accounting for merely 0.5% of available data^8^. This Eurocentric bias in GWAS results has led to disparities in PRS performance, with prediction accuracy in African people being about 4 folds lower than European people^1^. In response to these disparities, recent years have witnessed concerted global efforts to establish biobanks with enhanced diversity through the inclusion of non-European populations, such as All of Us^9^, China Kadoorie Biobank^10^, BioBank Japan^11^, GenomeIndia project^12^, Human Heredity and Health in Africa (H3Africa) Consortium^13^, and Uganda Genome Resource^14^. While these developments represent important steps forward, major regional and ancestry gaps remain.

While disparities among research participants and resources across ancestries and countries have been well studied, little is known about the representation and disparities of researchers themselves on the global stage. Among them, the American Society of Human Genetics (ASHG) and the European Society of Human Genetics (ESHG) conference^15,16^, as the two leading annual meetings in this field, have consistently demonstrated both important progress and ongoing challenges in human genetics. Over the past two decades, these conferences have maintained high academic standards and large-scale participation, mirroring the rapid advancements in human genetics research. While the genetic content presented at these meetings has been extensively studied^17^, little attention has been paid to the geographical distribution of their participants. This distribution may reflect persistent global disparities in human genetic resources and research, which in turn influence global equity in genomic medicine and healthcare.

To better understand the global disparities observed in human genetics research and resources, it is crucial to examine the representation of these leading platforms. Comparative analysis of regional representation at the two premier human genetics conferences, ASHG and ESHG, could provide valuable insights for promoting ancestral equity and reducing existing disparities. Although it has been shown that there are significant gender disparities in ASHG^18^, detailed studies into the geographical distribution of participants and longitudinal trends over the past two decades remain lacking.

Here, we conducted a comprehensive analysis by extracting author and institutional affiliations from conference abstracts published in ASHG and ESHG proceedings over the past 26 years. We identified the following four key findings: (1) abstract contributions have been heavily concentrated in North America and Europe; (2) participation from underrepresented continents like Africa and East Asia has grown but disparities persist; (3) an index of research visibility (oral presentations vs posters) shows particularly low visibility for underrepresented continents (Africa, parts of Asia, South America); and (4) gross domestic product (GDP) and Nature Index Share in biological sciences may be key contributing factors. Our findings provide critical insights into persistent imbalances in global participation and highlight underrepresented regions in the academic conferences of human genetics. We further discuss the potential implications of these disparities for the global human genetics community, emphasizing more broadly on equity, global health and genetics research, and diversifying research ecosystems. We also propose actionable strategies to foster more equitable international collaboration.

## Results

We first collected and analyzed all abstracts available from the official websites of the ASHG and the ESHG annual conferences. ASHG abstracts span from 1999 to 2024, while ESHG abstracts span from 2001 to 2024. Over these respective periods, ASHG presented a total of 77,448 abstracts across 26 years, averaging approximately 1,979 abstracts per year. In comparison, ESHG presented 43,823 abstracts over 24 years, with an annual average of approximately 1,826 abstracts (**Table S1**).

### Significant disparities in abstract presentations at continental and national levels

Significant regional differences are evident in abstract presentations to both the ASHG and ESHG conferences. At ASHG, North America dominated abstracts presentations, contributing 50,156 abstracts (64.8%) over the 26 years, with an average of 1,929 abstracts per year, exceeding the combined total abstracts from all other continents. This output was over 8 times higher than that of East Asia (6,048 abstracts, 7.8%, and 233 per year) and more than 80 times greater than that of Africa (598 abstracts, 0.8%, 23 per year) (**Fig. 1A-1B, Table S1**). As with ASHG, the dominance of European presentation at ESHG is more clear.

**Figure 1.**
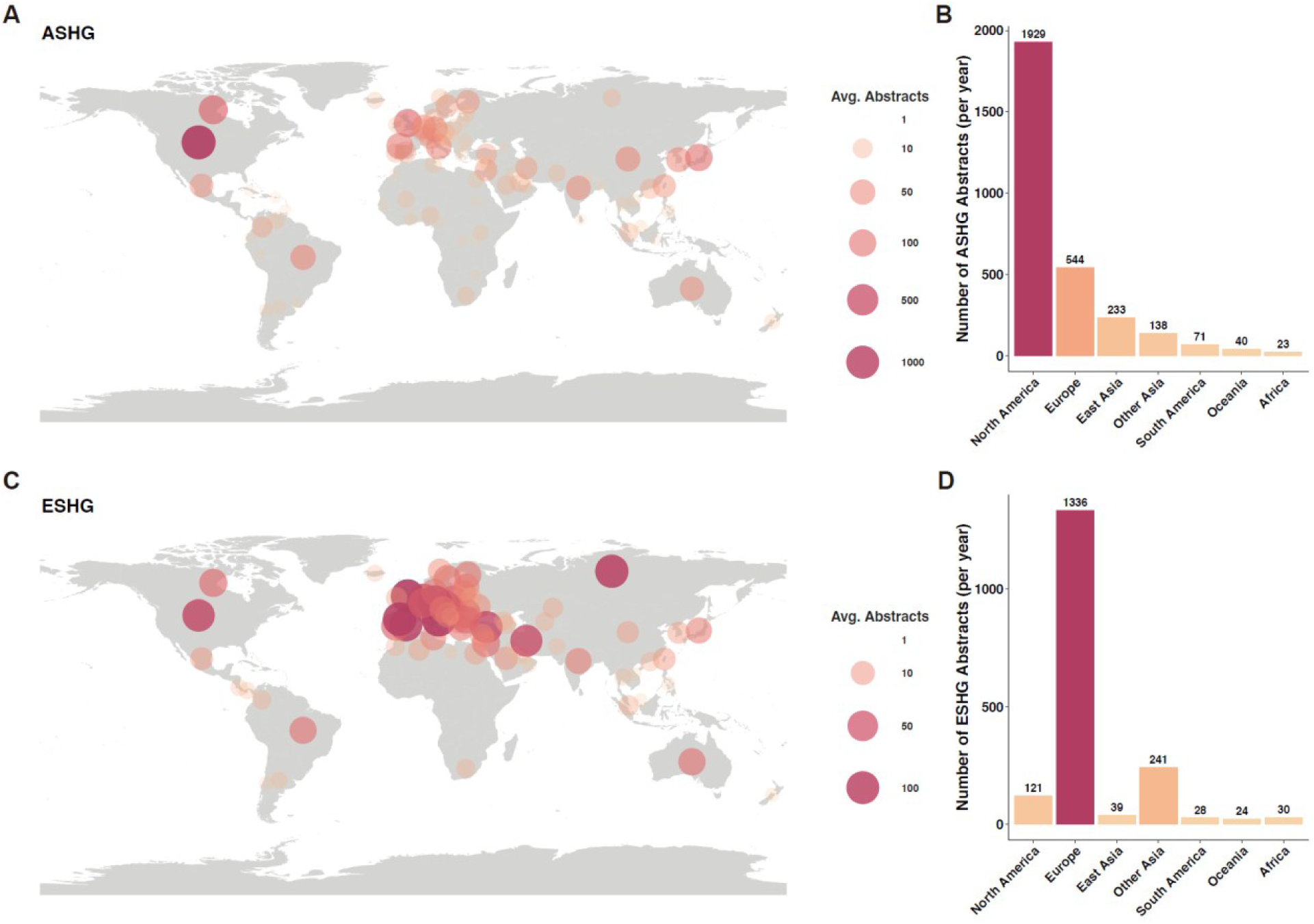
An overview of the average number of abstract presentations to ASHG and ESHG conferences. Figure A and B display the average annual number of abstracts presented at ASHG from 1999 to 2024, summarized by country (or region) and continent, respectively. Figure C and D present the corresponding data for ESHG from 2001 to 2024, also by country (or region) and continent.

European institutions led with 32,067 abstracts (73.2%, 1,336 per year), over 34 times that of East Asia (934 abstracts 2.2%, 39 per year) and 58 times that of Oceania, which had the fewest presentations (567 abstracts, 1.3%, 23 per year) (**Fig. 1C-1D, Table S1**). In addition, despite not being the host region, Europe ranked second in ASHG presentations, while North America ranked third in ESHG (**Table S1**).

We also analyzed the average number of abstracts at the country (or region) and sub-continent level. At ASHG, the United States alone contributed 1,711 abstracts, more than all other countries combined and 9.3 times that of the second-ranking country, Canada (**Fig. 1A, Table S2, Fig. S1**). In East Asia, Japan ranked fourth globally (n=106), exceeding abstracts from China (**Table S2, Fig. S1**). In South America, Brazil was the leading contributor, ranking eighth globally and far surpassing its continental peers, while in Africa, South Africa and Tunisia stood out (**Table S2**). Conversely, at ESHG, Germany had the highest average number of abstracts among all countries (138), but the gap among the top five countries was relatively small (**Fig. 1B, Table S2, Fig. S1**). While Europe dominated, the United States was the top non-European country (85, 8^th^) (**Table S2**). In Asia, Turkey and Iran had more abstracts than others, and in Africa, Tunisia and Egypt led in presentation counts (**Table S2**). Within each continent, certain sub-continents play a leading role at both meetings. For example, Southern and Northern Africa lead within the African continent, while Southern and Eastern Asia are prominent within Asia (**Fig. S2**).

Regional and national preferences for conference participation are evident. We compared the representation of the same continents and countries across both ASHG and ESHG conferences. The results indicate that the Americas and East Asia were more prominently represented at ASHG, whereas Europe, other parts of Asia (countries in Asia except for East Asia), and Africa showed a stronger presence at ESHG (**Fig. S3, Table S1**). At the country (or region) level, the United Kingdom exhibited a similar average number of abstracts at both ASHG and ESHG, even with a slight tilt toward ASHG, highlighting its central and dominant position in global human genetics research (**Fig. S4, Table S2**).

These results highlight substantial regional disparities in conference participation. Despite the heterogeneity observed at both country (or region) and sub-continent levels, North America and Europe consistently dominate ASHG and ESHG, regardless of whether they serve as host regions. In contrast, participation from Asia, Africa, and Oceania remains relatively limited.

### Stable trends in abstract presentations at ASHG and ESHG

To better understand the dynamics of global participation, we analyzed longitudinal trends in the number of abstracts presented at ASHG and ESHG. At ASHG, the number of abstracts increased modestly from 2,870 in 1999 to 2,945 in 2024, reflecting a 2.6% growth over 26 years (**Fig. 2A, Table S3**). Throughout this period, North America consistently contributed the highest share of abstracts, with an upward trend of 23.8% (**Fig. 2A, Table S3**). In contrast, the relative contributions from Europe, Oceania, and South America declined over the same period. Conversely, other continents showed some growth: abstract presentations from East Asia increased by 76.3%, from other parts of Asia by 74.7%, and from Africa by 327.3% (**Fig. 2A, Table S3**). At ESHG, the number of abstracts rose from 1,852 abstracts in 2001 to 2,793 in 2024, a 50.8% increase (**Fig. 2B, Table S4**). Over the past 24 years, participation grew across most continents, most significantly in Oceania (138.1%), other parts of Asia (101.1%), and Europe (72.5%) (**Fig. 2B, Table S4**). Compared to the relative stable trend in ASHG, ESHG has exhibited a clear upward trend. Nevertheless, the dominance and centre role of North America and Europe at both conferences has remained unchanged over time.

**Figure 2.**
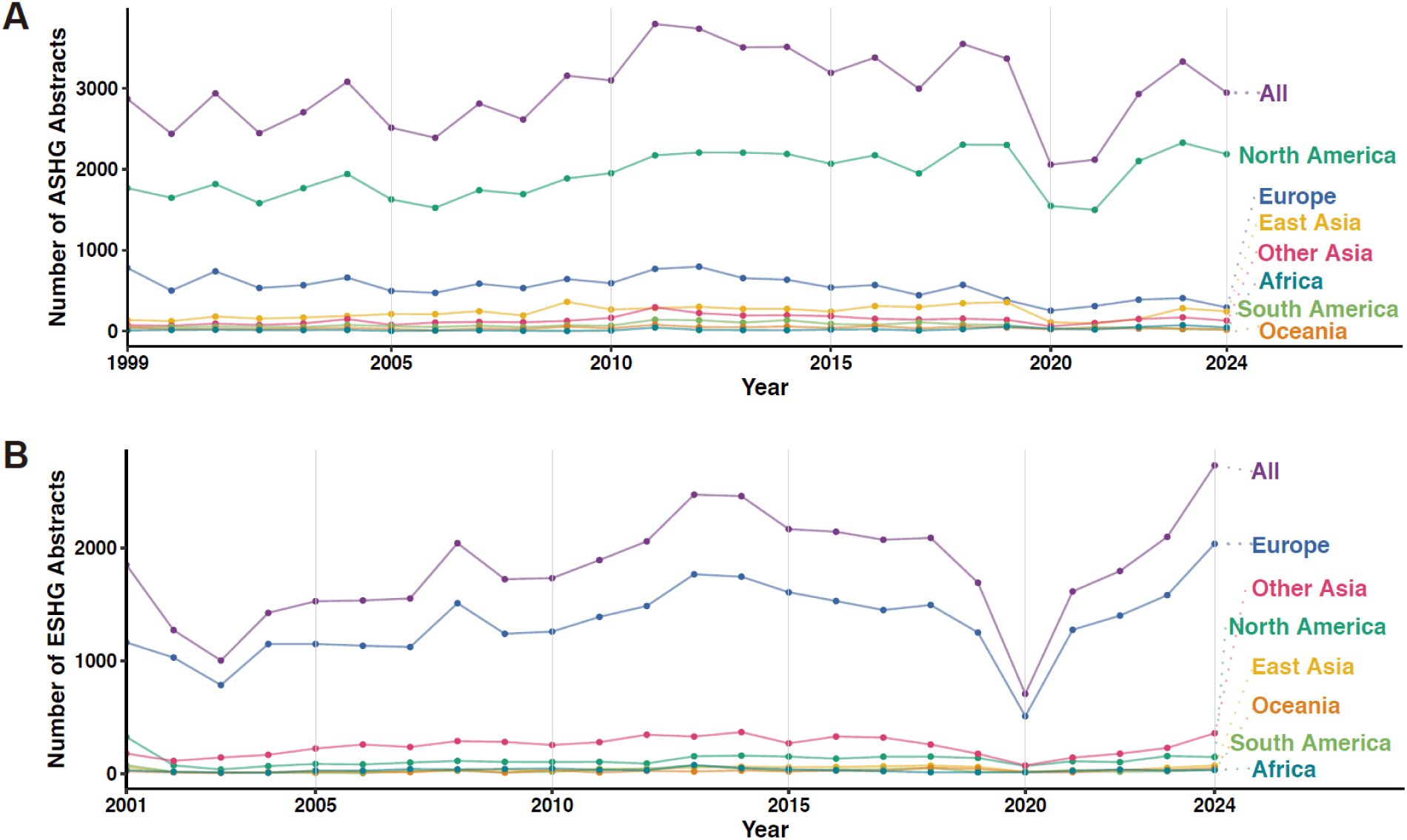
Continental trends in abstract presentations at ASHG and ESHG over the past two decades. Figure A shows the trend in the number of abstracts presented at ASHG by continents from 1999 to 2024. Figure B shows the trend in the number of abstracts presented at ESHG by continents from 2001 to 2024.

Country-level trends further underscore the persistent global imbalance at ASHG and ESHG. South Africa, the most represented African country at ASHG, presented only 0-16 abstracts annually, while the United States consistently led with 1,500-2,000, far surpassing all other countries (**Fig. 3A**). Brazil and Australia dominated in South America and Oceania, respectively (**Fig. 3A**). In East Asia, Japan contributed the highest number of abstracts, followed by China and South Korea (**Table S2**). In addition, the number of abstracts in China rose steadily before 2020, from ∼20 to ∼100 per year. A similar pattern is observed at ESHG, with a few countries, such as the United States, Australia, and Brazil, remaining top contributors in their continents (**Fig. 3B**). Most other countries contributed fewer abstracts, but their abstracts counts showed modest growth, such as Tunisia and Turkey (**Fig. 3B**). In East Asia, Japan led consistently, while China ranked fourth (**Table S2**). Among European countries, annual abstract counts were relatively balanced, generally fluctuating between 100 and 400 (**Fig. 3B**).

**Figure 3.**
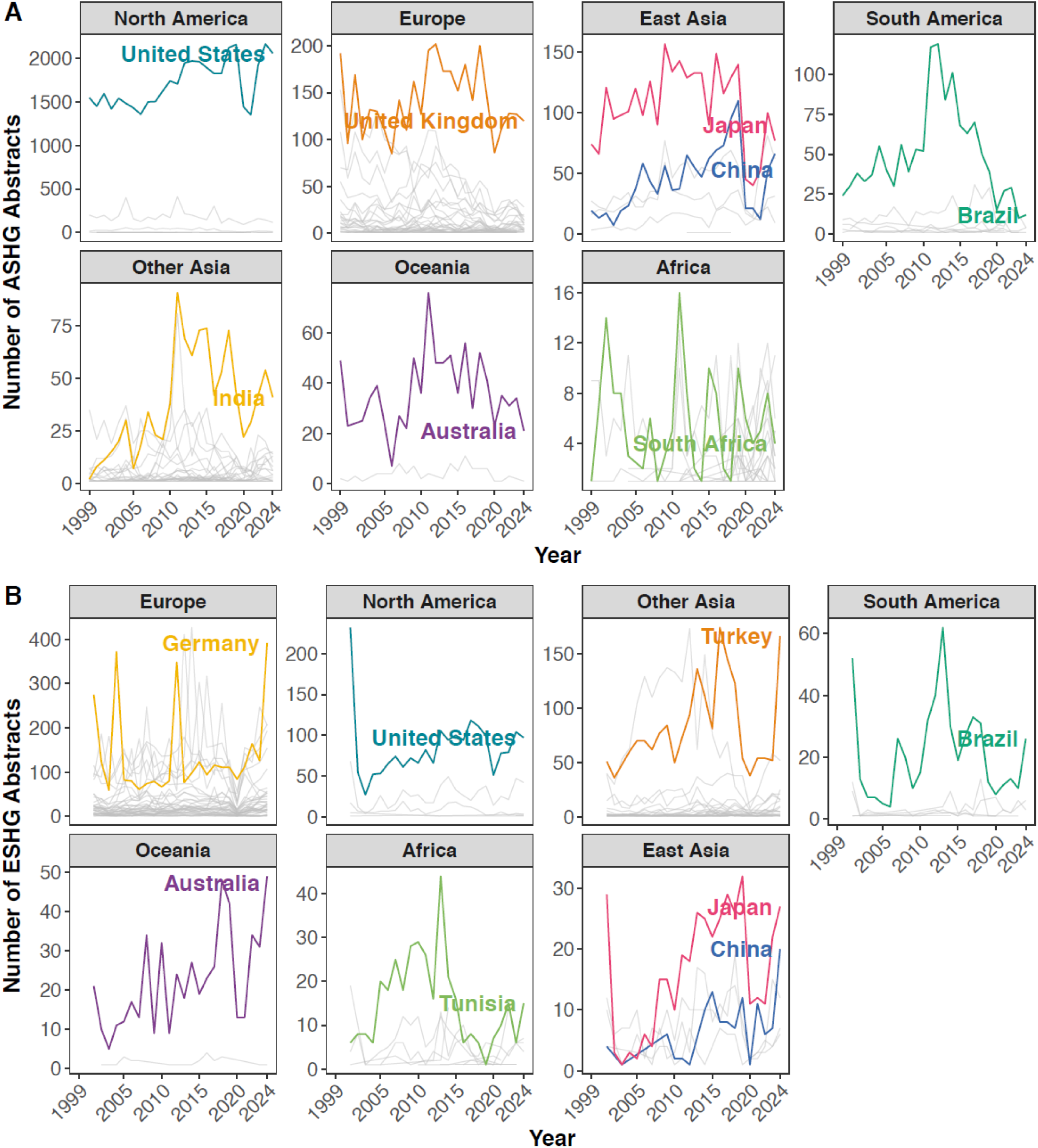
Country(or region)-level trends in abstract presentations at ASHG and ESHG over the past two decades. Figure A shows the trends in the number of abstracts presented by individual country (or regions) across each continent at ASHG. Figure B shows the trends in the number of abstracts presented by individual countries (or regions) across each continent at ESHG. In both figures, the country (or region) with the highest number of abstracts within each continent are highlighted in color, while other countries (or regions) are shown in gray. In the East Asia continent, both Japan (the highest) and China are specifically highlighted.

Collectively, over the past two decades, ASHG and ESHG have maintained stable abstract volumes, with North America and Europe consistently dominating. Although countries in East Asia, Africa, and other Asian continents have shown increased participation, abstract contributions remain concentrated in a few nations (or regions) within each continent.

### Disparities in oral versus poster presentations

We categorized all ASHG and ESHG abstracts into two types, oral presentations and posters, and compared the regional proportion across both types. Consistent with the results above, the majority of oral presentations were from North America and Europe (**Figs. 4A-B, Table S5**). At ASHG, North America accounted for 73.5% of oral presentations, followed by Europe (20.7%), both significantly higher than East Asia (2.4%) and South America (0.1%). At ESHG, Europe contributed 77.1% of oral presentations, followed by North America (15.0%), both also significantly exceeding the counts from East Asia (1.1%) and South America (0.2%). In addition, considering the relative proportion of these two formats, Oceania showed a higher proportion of oral presentations, while Asia, Africa, and South America contributed more posters in each continent (**Figs. 4A-B, Table S5**).

**Figure 4.**
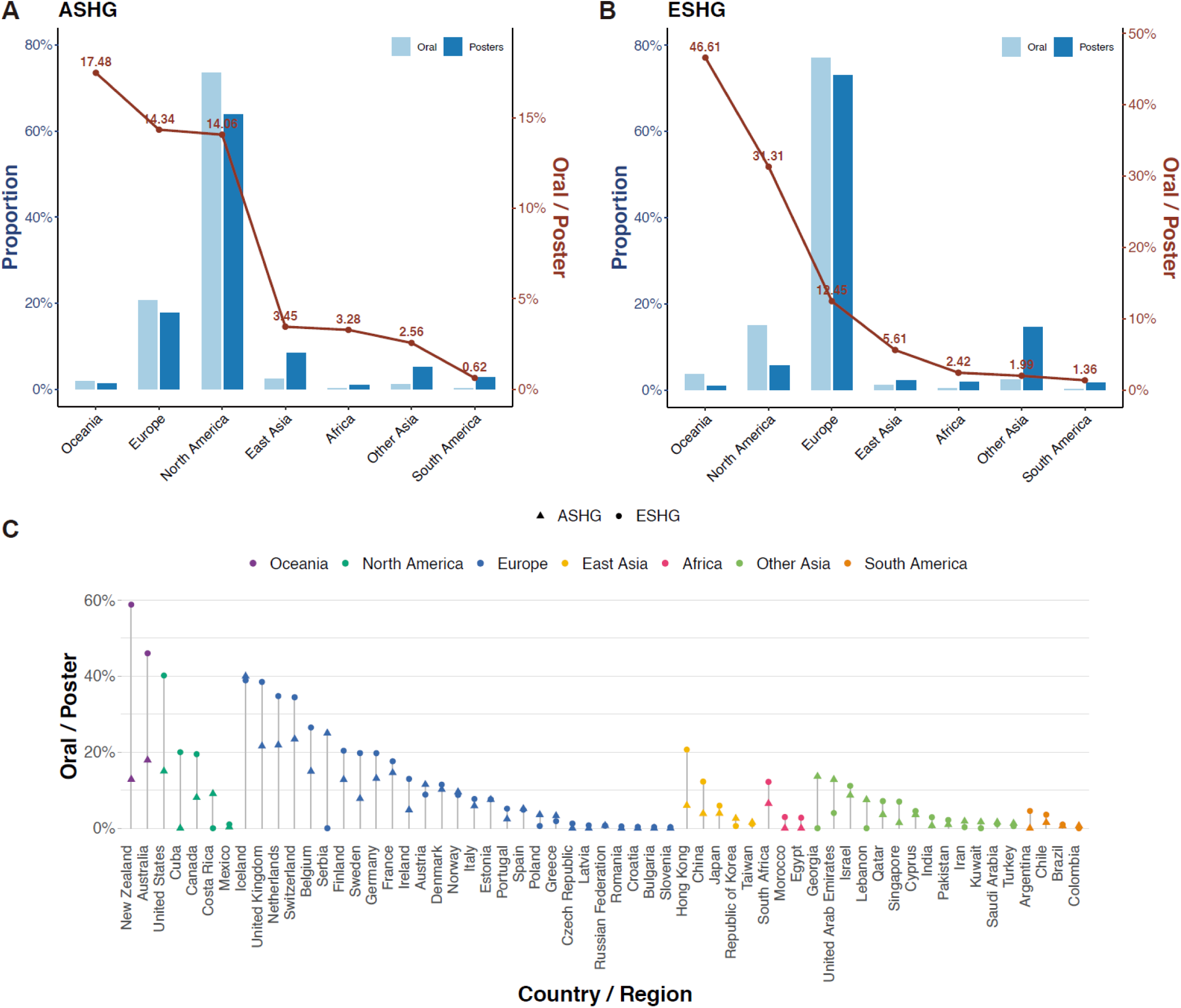
Continental and national distribution and oral-to-poster ratios at ASHG and ESHG. Figure A displays the proportion of oral presentations and posters contributed by each continent at ASHG (left panel), and the ratio of oral presentations to posters (oral/poster) for each continent (right panel). Figure B shows the same for ESHG. Figure C presents the oral-to-poster ratio for countries (or regions) that contributed more than five total abstracts and had non-zero ratios in both conferences.

To benchmark the possibility of oral presentations, we calculated the ratio between the number of oral presentations and posters within each continent (**Methods**). In both conferences, this ratio exceeded 10% in North America, Europe, and Oceania, suggesting a greater likelihood of selection for oral presentations, while it remained below 6% in Asia, Africa, and South America (**Figs. 4A-B, Table S5**). For example, in the ASHG, the oral-to-poster ratio was 14.06% for North America, 3.45% for East Asia, and just 0.62% for South America, indicating that abstracts from North America were approximately 4 times more likely to be selected for oral presentation than those from East Asia, and 23 times more likely than those from South America. In the ESHG, the ratio was 12.45% for Europe, 5.61% for East Asia, and 1.36% for South America, meaning that abstracts from Europe were about 2 times more likely to be selected than those from East Asia, and nearly 9 times more likely than those from South America. Notably, Oceania exhibited the highest oral-to-poster ratio in both conferences, with values of 17.48% in ASHG and 46.61% in ESHG. These ratios were 5 and 8 times higher than East Asia, and 29 and 33 times higher than South America, respectively. In addition, Europe ranked second at ASHG with an oral-to-poster ratio of 14.34%, while North America ranked second at ESHG with a ratio of 31.31%. Although these ratios are influenced by the total number of abstracts submitted from each continent, they suggest that abstracts from these three continents are more likely to be recognized as higher quality or higher impact. In comparison, although East Asia had higher participation, its oral-to-poster ratio was lower. Africa faced an even lower ratio alongside limited abstract presentations, further reducing its visibility (**Figs. 4A-B, Table S5**).

We also calculated the oral-to-poster ratio for each country (or region) (excluding countries with fewer than five total abstracts across both conferences). In ASHG, the highest oral-to-poster ratio was observed in Iceland (40.0%), followed by Switzerland (23.44%), the Netherlands (21.88%), the United Kingdom (21.60%), and Australia (17.92%) (**Fig. 4C, Table S6**). Additionally, Uganda (15.79%) also exhibited relatively high ratios. In contrast, East Asian countries or regions such as Hong Kong (China, 6.0%), Japan (3.89%), and China (3.83%) all had ratios below 6.0% (**Table S6**). In ESHG, New Zealand (58.82%) and Australia (46.02%) had the highest ratios, followed by the United States (40.18%), Luxembourg (40.00%), and Iceland (38.89%) (**Fig. 4C, Table S6**). Among East Asian continents, Hong Kong (China) had the highest ratio (20.69%), followed by China (12.26%). In Africa, only South Africa had a ratio greater than 10% (12.20%). Besides, we also compared the ratios between different countries based on whether English is an official language. Countries where English is an official language had higher ratios than other countries (**Fig. S5**).

The analysis of presentation types shows that North America and Europe not only lead in total abstract counts but also have higher chances of oral presentations, a format generally associated with higher research visibility. In contrast, continents such as East Asia, Africa, and South America are underrepresented in oral sessions, despite increasing poster participation. Oceania, particularly New Zealand and Australia, stands out with higher oral-to-poster ratios at both ASHG and ESHG, even though its modest total abstract count. In addition, a few countries like Iceland, Luxembourg, and Uganda, also achieved high ratios, although overall visibility from underrepresented continents remains limited.

### Potential causes of regional differences in conference participants

Finally, we explore potential national-level factors underlying the observed regional disparities in abstract and oral presentation contributions. We collected data on six indicators: population size, gross domestic product (GDP), number of researchers, education index, gross domestic expenditure on research and development (GERD), and Nature Index Share in biological science (Share, which indirectly reflects the high-quality research capabilities of different countries in the field of biological sciences). These variables were used to construct multivariable linear regression models to examine their associations with the average number of total abstracts and oral presentations per country (or region). After applying natural log transformation (logₑ) and standardization, most variables approximated a normal distribution, and the absolute pairwise correlations among these six variables ranged from 0.034 to 0.804 (**Fig. S7**). Following variable transformation and stepwise model selection, we obtained four final regression models (**Tables S7–S8**). When modeling the average number of abstracts as the dependent variable, we found that GDP was a significant predictor in both ASHG and ESHG (**Table S7**). When modeling the average number of oral presentations, Nature Index Share was a significant factor in both ASHG and ESHG (**Table S8**). In the models for two types of abstract average counts, the standardized regression coefficients (per SD) for GDP were moderate (ASHG: 0.37; ESHG: 0.35). While, in the models for oral presentations, the coefficients (per SD) for Nature Index Share were higher (ASHG: 0.82; ESHG: 1.03), indicating that a one-standard-deviation increase in Nature Index Share was associated with an increase of more than 0.8 oral presentations.

In addition, we incorporated two categorical variables into the models to assess the influence of the continent and official language, respectively. In the continent-based models, the Nature Index Share remained a significant predictor of the average number of oral presentations (**Tables S9-10**), suggesting it is a stable factor associated with research visibility. After adjusting for GERD, the number of researchers, and Nature Index Share, differences in oral presentation counts between Europe and other continents became small, such as North America, Oceania, other parts of Asia, and Africa (**Table S10**). In the language-based models, after controlling for other national-level factors, there was no significant difference in oral presentation counts between countries that do and do not list English as an official language (**Tables S11-12**).

We examined the relationship between national GDP and the average number of abstracts, as well as Nature Index Share and the average number of oral presentations, and additionally calculated the ratio of average abstracts per log_₁₀_(GDP) and oral presentations per log_₁₀_(Nature Index Share) (**Fig. 5**). At ASHG, the United States, Canada, and the United Kingdom had the highest average numbers of abstracts and oral presentations relative to their log-transformed GDP and Nature Index Share values (**Fig. 5A and Fig. 5C**). At ESHG, Italy, Germany, and the United Kingdom led in average abstracts per log_₁₀_(GDP) (**Fig. 5B**), while the United Kingdom, Netherlands, and the United States ranked highest in oral presentations per log_₁₀_(Nature Index Share) (**Fig. 5D**). Despite its high GDP and substantial contributions to the Nature Index Share, China ranked lower in both conferences across all metrics (**Fig. 5**).

**Figure 5.**
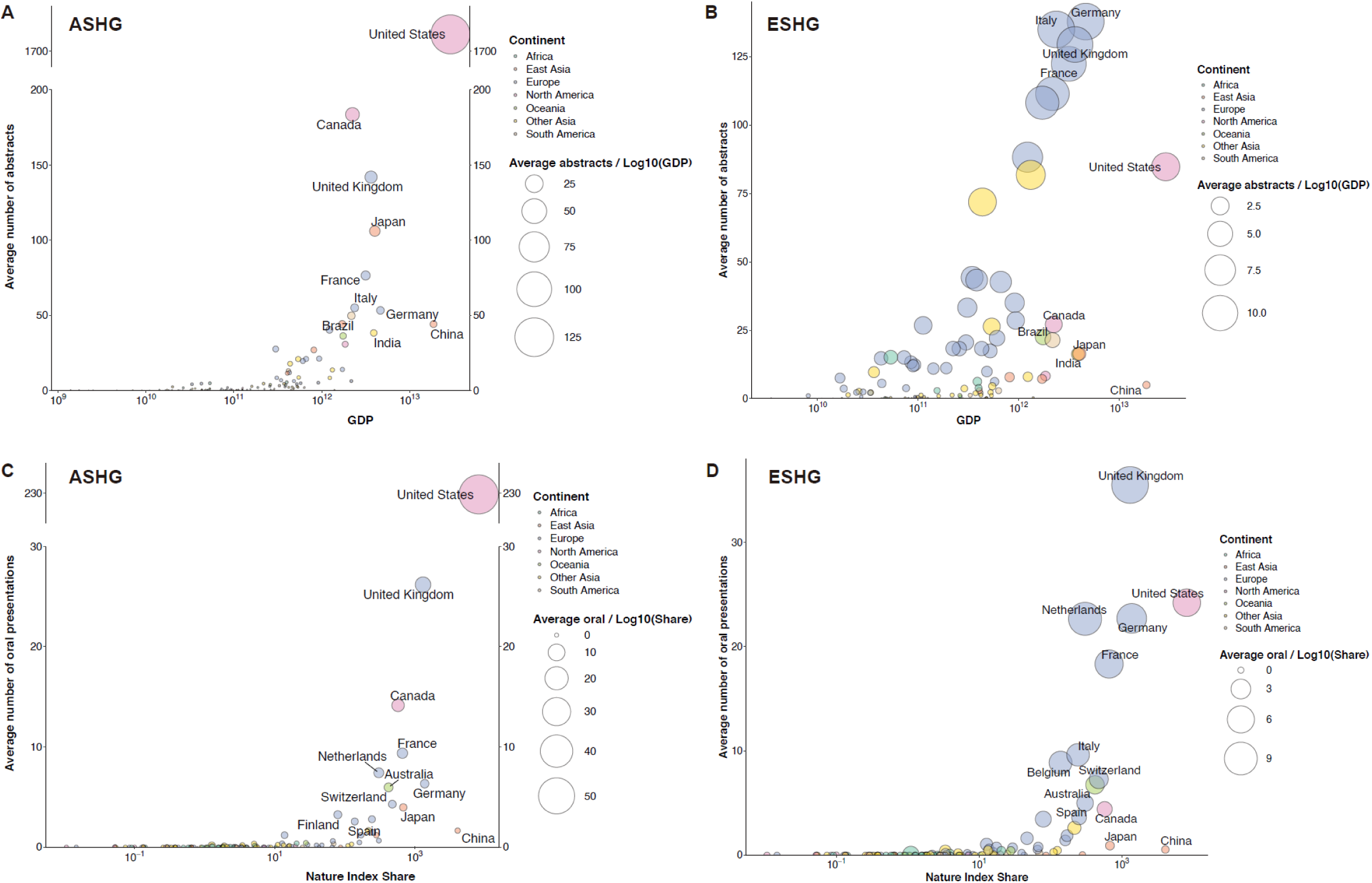
National GDP and Nature Index Share and the average of presented abstracts in ASHG and ESHG. Panel A displays each country’s (or region’s) GDP, average number of abstracts, and average abstracts per log₁₀(GDP) at the ASHG conference. Panel B shows the corresponding data for the ESHG conference. Panels C and D present each country’s (or region’s) Nature Index Share in biological science, average number of oral presentations, and average oral presentations per log₁₀(Share) at ASHG and ESHG, respectively. Dot colors represent different global continents, while dot sizes reflect the average number of abstracts per log₁₀(GDP) or oral presentations per log₁₀(Share). The top ten countries (or regions) by GDP, Nature Index Share, and average number of abstracts are labeled in each panel. GDP: gross domestic product (in US dollars). Share: Nature Index Share in biological science.

In summary, our findings suggest that national-level indicators, including GDP and Nature Index Share in biological sciences, contribute to the observed disparities in abstract and oral presentation counts across countries. In addition, Nature Index Share was significantly associated with the quality of academic contributions, as reflected by a higher likelihood of abstracts being selected for oral presentations.

## Discussion

Regional disparities remain a persistent challenge in human genetics research, as reflected in the participation at the two leading global conferences, ASHG and ESHG. In this study, we systematically analyzed the geographic distribution of abstracts presented at these conferences over the past two decades. Our findings reveal consistent continental imbalances: regardless of the host region, North America and Europe contributed the overwhelming majority of abstracts, while continents such as Africa, Oceania, and East Asia were significantly underrepresented. Trend analyses showed that these disparities have persisted throughout the 26-year period, with little sign of narrowing. Additionally, North America and Europe also dominated the oral presentation part, further indicating their central position in the global genetic research. Moreover, under-represented continents (Africa, parts of Asia, South America) had a very low “visibility” by calculating an oral-to-poster ratio index. However, Oceania stood out with the highest oral-to-poster presentation ratio, reflecting greater research visibility despite lower overall output. GDP and Nature Index Share, representing the economic and scientific capacity of a country (or region), respectively, were identified as key determinants of the average number of abstracts and oral presentations. These results highlight the ongoing disparities in both research output and global visibility within human genetics conferences.

Regional disparities have long persisted in human genetics research. Eurocentric biases are still common in genetic research^19^, and our analysis confirms that from the angle of representation of human geneticists at premier conferences. This is particularly concerning given that some of the most underrepresented continents, such as Africa and Asia, harbor the highest levels of genetic diversity and, in the case of Asia, the largest populations globally^20^. Although recent efforts have aimed to improve global diversity, such as the development of regional biobanks and multi-ancestry studies, the pace of change has been slow. Studies that include participants from underrepresented regions or multiple ancestries have only marginally increased^21,22^. Consistent with this, our findings show that only 18% of all abstracts at ASHG and ESHG over the past two decades originated from outside North America and Europe. This persistent pattern not only reflects issues of inequity but also limits scientific discovery. Underrepresentation in genomic research can result in missed opportunities to identify shared or population-specific disease-causing variants, restrict the generalizability of polygenic risk scores, and hinder the development of globally representative whole genomes sequencing^23–25^. Therefore, addressing these disparities is not only a matter of fairness, but a scientific urgency.

Oral presentations at academic conferences offer the visibility for research findings and provide valuable opportunities for scientists, particularly those from underrepresented continents and countries (or regions), to receive feedback, engage in broader discussions, and connect with the global research community. However, our analysis suggests that access to this opportunity is disproportionate. North America and Europe had the highest proportions of oral presentations, consistent with the persistent disparities in human genetics. Oceania, particularly Australia, showed the highest likelihood of oral presentations, despite a smaller total number of abstracts, indicating its strong position in the field. Although many biobanks have been established in the non-European population or designed to include diverse ancestries, their representation in both oral and poster presentations at conferences remains relatively low. For instance, although East Asia ranked third overall, Japan (3.89% at ASHG; 5.93% at ESHG) and China (3.83% at ASHG; 12.26% at ESHG) had low visibility. Notably, China presented only 36 oral abstracts at ASHG and 13 at ESHG over the past 26 years. African and South American countries performed even less favorably, with fewer than 10 oral presentations each over the past two decades. Their low oral-to-poster ratios further reflect reduced visibility and limited opportunities to showcase their work on prominent platforms.

Structural advantages including technological advances, research infrastructure, and funding play a key role in genome research^21^. In this study, we examined six national level variables and two categorical factors to explore their impact on abstract presentations at academic conferences. Among them, GDP, a standard indicator of a country’s economic strength, showed a strong positive association with the average number of abstracts presented. This aligns with the reality that large-scale genetic studies are costly, and many developing countries lack the financial resources to support such research. For oral presentation counts specifically, we found the Nature Index Share in biological sciences had the strongest and most consistent association. The Nature Index tracks contributions to high-impact journals, and the Share metric reflects both authorship proportion and collaboration scale at the country or continental level^26^. We used this as a proxy for high-quality scientific output. Since oral presentations often reflect higher research quality, countries or regions with higher Nature Index Share were more likely to be selected for talks. Importantly, GDP and Nature Index Share were highly correlated (r > 0.7), highlighting their central roles in both quantity and quality of human genetics research.

To address these disparities, several steps are essential. For conference organizers, first, organisers should provide dedicated resources—such as multilingual virtual workshops on abstract preparation—to broaden the pool of high-quality submissions from under-represented regions. Second, oral-presentation slots could be allocated to a higher proportion to under-represented regions, with an additional equity buffer to ensure visibility in high-profile sessions. Complementary measures—including hybrid presentation options, travel bursaries and accommodation support—would further lower financial and logistical barriers. Finally, systematic bias-mitigation initiatives (blind scoring, unconscious-bias training and yearly diversity reports) can foster a more inclusive global genetics community. More broadly, continued global efforts are needed, including sufficient government funding, better infrastructure, and stronger international collaborations^21^.

This study has several limitations that should be acknowledged. First, due to inconsistent formatting and incomplete metadata in some publicly available conferences records, the extraction of institutional affiliation was occasionally challenging; however, these instances were rare (< 0.2%). Moreover, given that many of the findings presented at ASHG and ESHG are subsequently published in peer-reviewed journals, such as *The American Journal of Human Genetics* and *European Journal of Human Genetics*, our study instead focused on the geographic and institutional aspects of global participation rather than scientific merit. In addition, in our multivariable regression model, we used Nature Index Share as a proxy for research quality, as it reflects timely contributions to high-impact publications in natural and health sciences. Despite these limitations, this study offers important insights into the structural and geographic disparities in global human genetics research.

In conclusion, our study underscores the ongoing structural inequities in global engagement with human genetics research. By recognizing and actively mitigating these gaps, the human genetics community can move toward a more inclusive and globally representative future, which will ultimately accelerate discoveries and benefit all populations.

## Methods

### Data sources

We accessed publicly available abstracts from the annual meetings of the American Society of Human Genetics (ASHG) and the European Society of Human Genetics (ESHG), spanning over two decades. ASHG abstracts were obtained from: https://www.ashg.org/meetings/future-past/abstract-archive/ and covered the years 1999 to 2024^15^, while the ESHG abstracts from 2001 to 2024 were collected from https://www.eshg.org/conferences/past-eshg-meetings^16^. All abstracts were categorized as either oral presentations or posters (abstracts from ASHG 2005 to 2008, which lacked presentation type information, were excluded from the following oral or poster specific analysis). In ASHG, oral presentations included Platform and Plenary sessions. In ESHG, oral presentations comprised ESHG Spoken Presentations and EMPAG Spoken Presentations, while posters included Electronic Posters, Posters, EMPAG Posters, and Published Abstracts.

All abstracts were processed in PDF format. We used the Python package pdfminer.six (version 20221105) to extract text from each file in batch mode. For each abstract, we retrieved data including the title, author names, and first-listed institutional affiliation. Country (or region) and continent assignments were determined based on the first-listed institution. If the country could not be identified from the first institution, we sequentially reviewed subsequent affiliations until a clear country assignment could be made. Abstracts were excluded if no institutional information was provided or if country determination was not possible.

Geospatial data for each country (or region) were obtained using the ‘ne_countries’ function from the R package rnaturalearth (version 1.0.1), based on Natural Earth vector data v4.1.0 (March 2018)^27^. The geographic centroid coordinates (latitude and longitude) of each country (or region) were retrieved using the R package sf (version 1.0-16). Countries (or regions) were grouped into seven major world continents: North America, South America, Europe, Africa, East Asia, Other parts of Asia (All Asian countries/regions outside East Asia), and Oceania.

### Ratio of oral presentations to posters by region

To assess the relative likelihood of abstracts being selected as oral presentations across different regions, we calculated a standardized ratio:

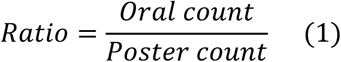

In this formula, “Oral count” represents the number of oral presentations from each continent or country (or region), while “Poster count” refers to the number of posters from each continent or country (or region).

### Multivariable regression analysis

To investigate the factors influencing the average number of abstracts and oral presentations presented from each country (or region), we constructed multivariable linear regression models using six national-level indicators. Country-level population size and gross domestic product (GDP) data were obtained from the World Bank Open Data^28^. Data on gross domestic expenditure on research and development (GERD, as % of GDP) and number of researchers in each country (or region) were sourced from the UNESCO Institute for Statistics^29^, and education index data were retrieved from World Population Review^30^. The Nature Index Share in biological sciences was obtained from the country (or region) tables on the Nature Index website^26^. In addition, we included two categorical variables, continent and language, in these models. Information on official languages was extracted from the corresponding entry on Wikipedia^31^. If English was among the official languages of a country (or region), it was classified as a country (or region) with English as an official language. The latest year of data was selected for analysis. Pairwise relationships among all variables were visualized using the *ggpair* function from the GGally package (version 2.2.1) in R^32^.

We selected the average number of abstracts and the average number of oral presentations per country (region) for each of the two meetings as the dependent variables and six national-level indicators, population size, GDP, GERD, number of researchers, education index, and Nature Index Share as the independent variables. For the two categorical variables, we included them in the models separately. For the continent, North America was used as the reference category in ASHG analyses, while Europe served as the reference in ESHG analyses. For language, countries with English as an official language were used as the reference group. All quantitative variables were transformed using the natural logarithm (logₑ) and subsequently standardized. To impute the missing value, we applied multiple imputation via predictive mean matching (PMM) using the R package mice (version 3.16.0). Based on these datasets, we fitted 12 multivariable linear regression models corresponding to the combinations of dependent and independent variables. The initial ASHG models are as follows:

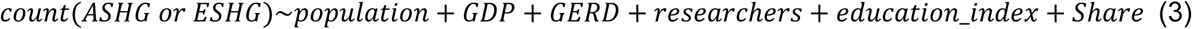

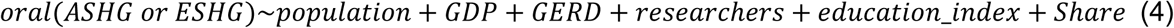

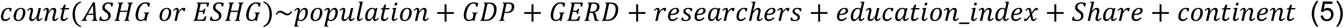

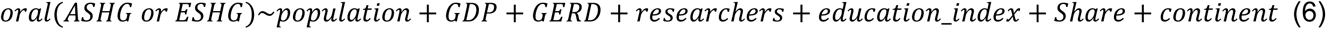

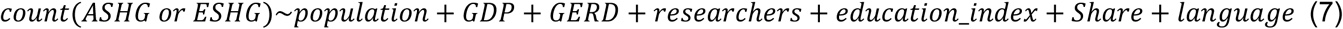

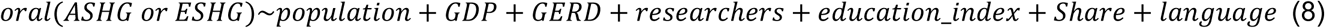

In these models, *count(ASHG or ESHG)* is the average number of abstracts presented in ASHG or ESHG for each country (or region), and *oral(ASHG or ESHG)* is the average number of oral presentations in ASHG or ESHG for each country (or region).

In all multivariable regression models, except for the two categorical variables, continent and language, all independent and dependent variables were log-transformed and standardized.

After that, we performed stepwise selection based on Akaike’s Information Criterion (AIC) with stepAIC from the R package MASS (version 7.3-58.2). The resulting analysis of deviance table and final model summary were inspected to identify the variables retained in the optimal model and to evaluate overall fit. In the multivariable regression models, independent variables with p-values < 0.05 were considered statistically significant.

### Statistical analysis

To compare the proportion of abstracts from each continent between the two conferences, we applied the two-proportion z-test. For comparisons among sub-continents, we used the Wilcoxon test. A p-value < 0.05 was considered statistically significant. Unless specified otherwise, all statistical analyses and figures were conducted using R (version 4.2.3).

## Data Availability

All data generated or analyzed in this study are publicly available.

## Supplementary Figures

**Supplementary Figure 1.**
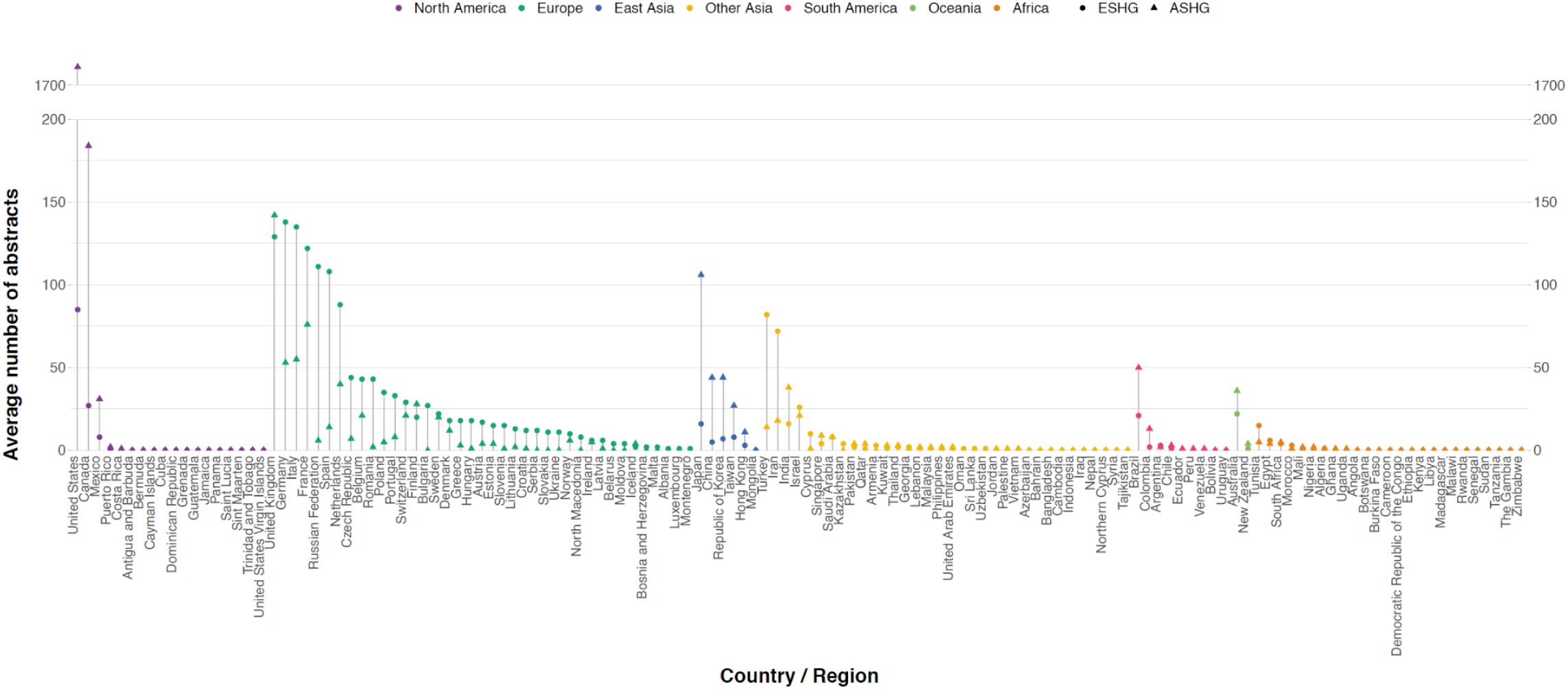
The average number of abstract presentations to ASHG and ESHG conferences. This panel shows the average annual number of abstracts presented at the ASHG and ESHG conferences. Point shape denotes the conference, and color indicates the continent. The y-axis represents the average number of abstracts per country or region. We compared the average number of abstracts presented in each country (or region) across both meetings. We found countries (or regions) in North America and Europe had higher averages, while South America, Oceania, and Africa had lower counts. In East Asia, Japan exceeded China in average abstract numbers.

**Supplementary Figure 2.**
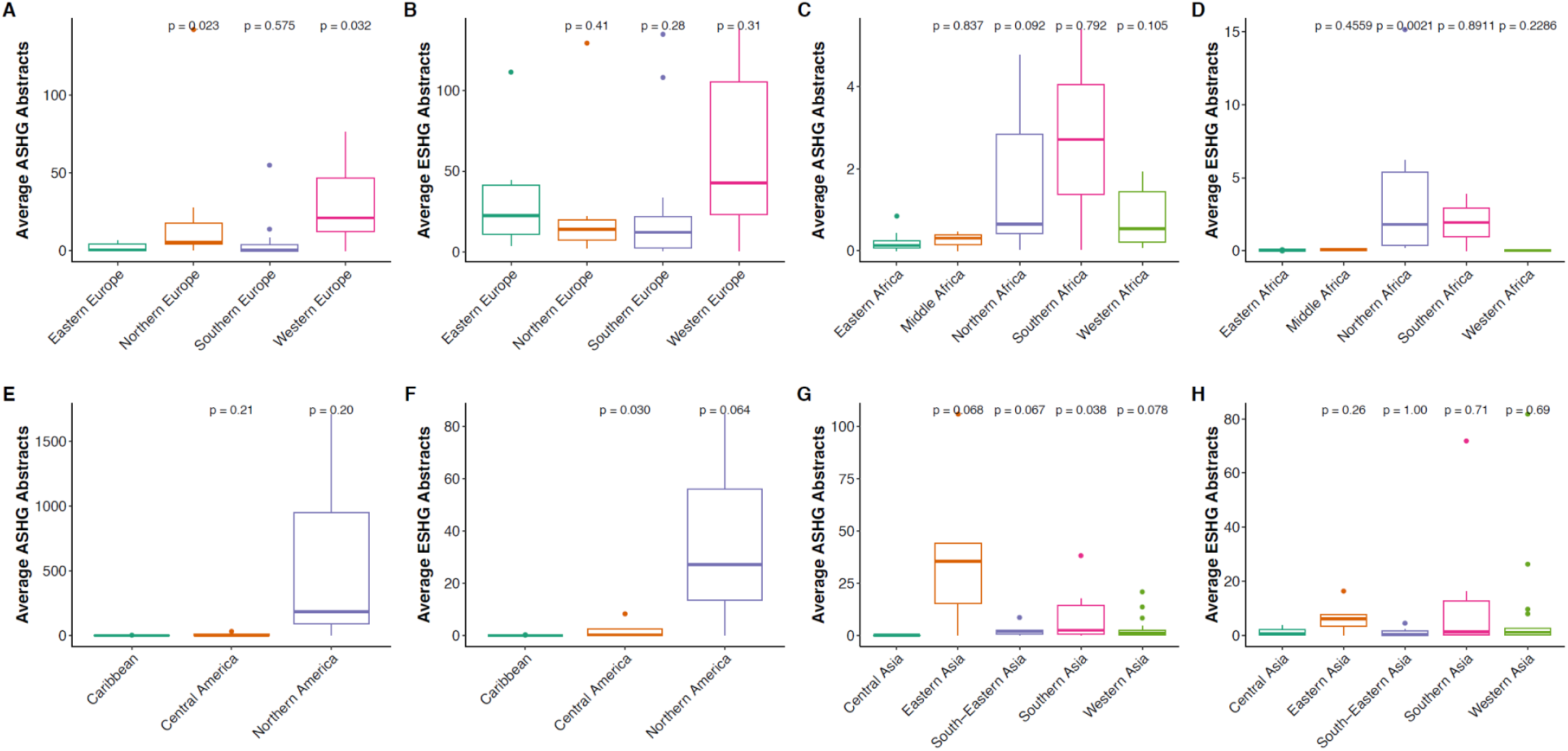
Sub-continental comparison of average abstract presentations within continents. The boxplots show the distribution of the average number of abstracts presented across sub-continents within each continent. In each panel, the first sub-continent is the reference group. Wilcoxon tests were conducted to assess whether the differences in average abstract counts between sub-continents are statistically significant. We compared the distribution of the average number of abstracts across sub-continents within each continent and observed notable intra-continental heterogeneity. For instance, within Asia, East Asia contributed a higher number of abstracts; in Europe, Western Europe led in abstract counts; and in Africa, both North Africa and South Africa showed higher abstract counts compared to other sub-continents.

**Supplementary Figure 3.**
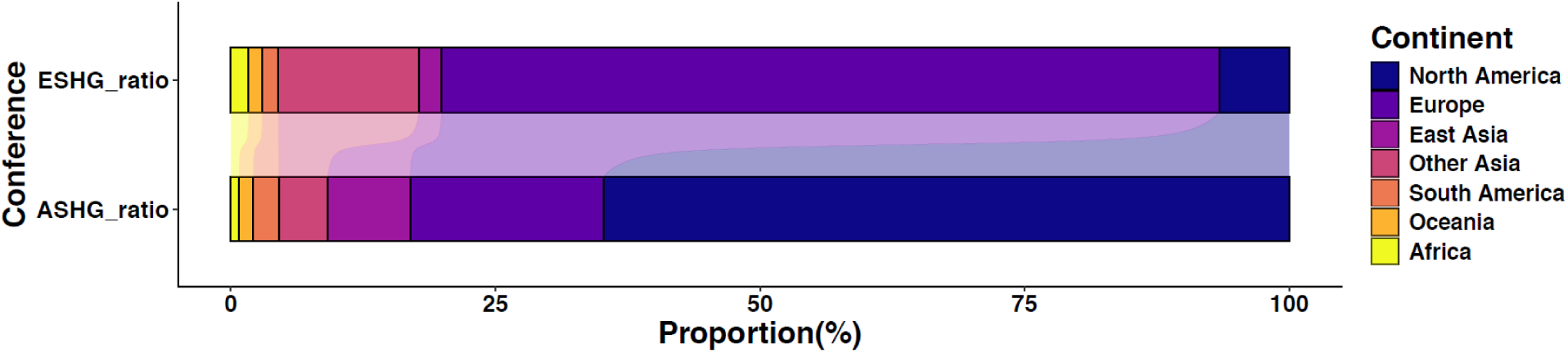
The proportion of each continent presenting abstracts at ASHG and ESHG. Figure shows the proportion of abstracts by different continents in these two conferences. We calculated the proportion of abstracts presented by each continent in the ASHG and ESHG. The results indicate that Europe, other parts of Asia, and Africa showed a stronger presence at ESHG, whereas the Americas, East Asia, and Oceania were more prominently represented at ASHG.

**Supplementary Figure 4.**
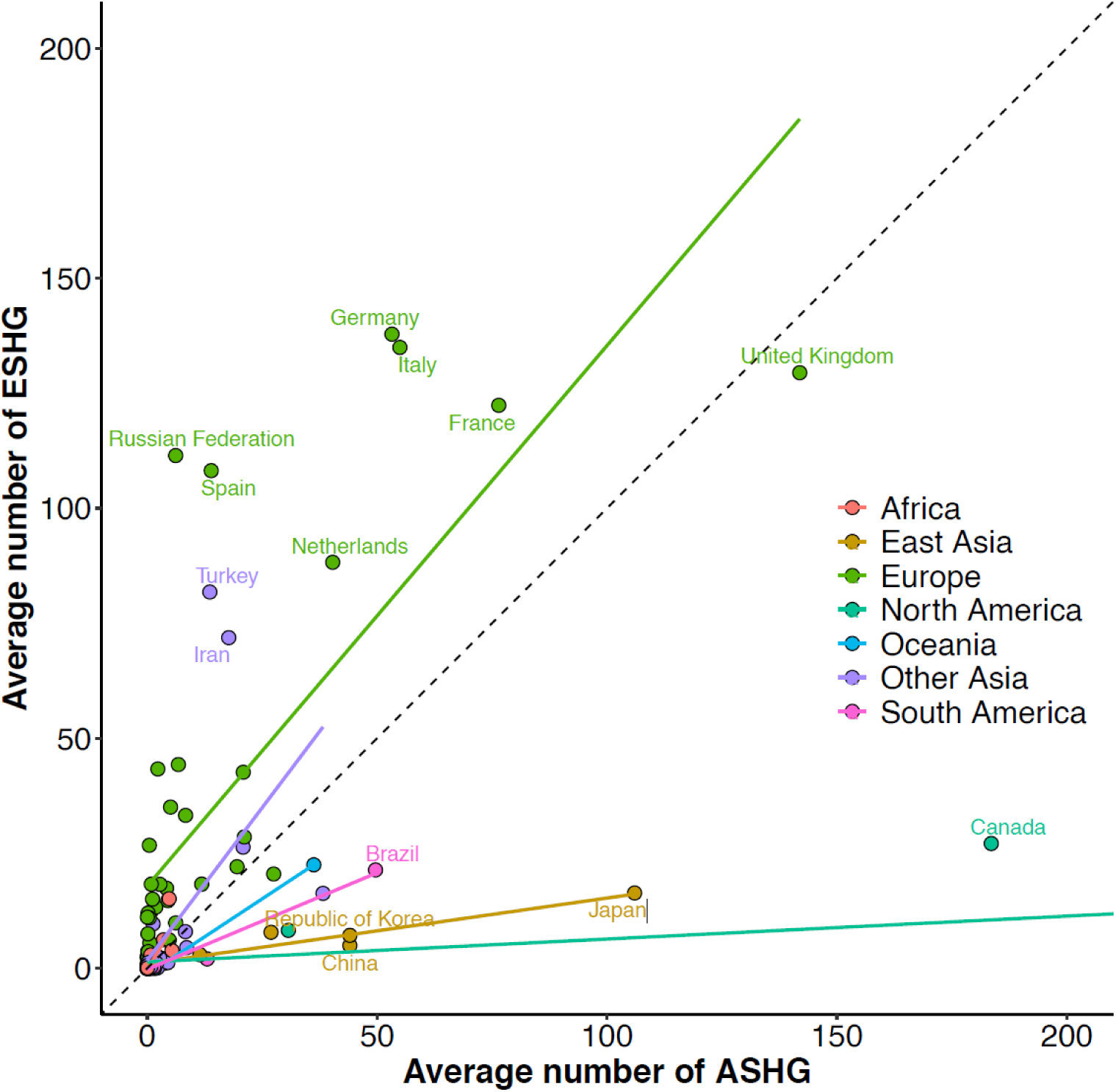
The average number of abstracts presented in each country (or region) at ASHG and ESHG. The scatter plot shows the average number of abstracts submitted by each country/region to the ASHG and ESHG conferences. The dashed diagonal line represents the identity line (y = x). Colored solid lines show continent-specific linear fits. To improve visualization, the **United States**, an extreme outlier with coordinates (**1711.38, 84.75**), is excluded from the plot due to its disproportionately high ASHG abstract count. We compared country-level participation and observed distinct continental tendencies in these two conferences. The United Kingdom showed comparable levels of participation in both ASHG and ESHG, with a slight preference for ASHG.

**Supplementary Figure 5.**
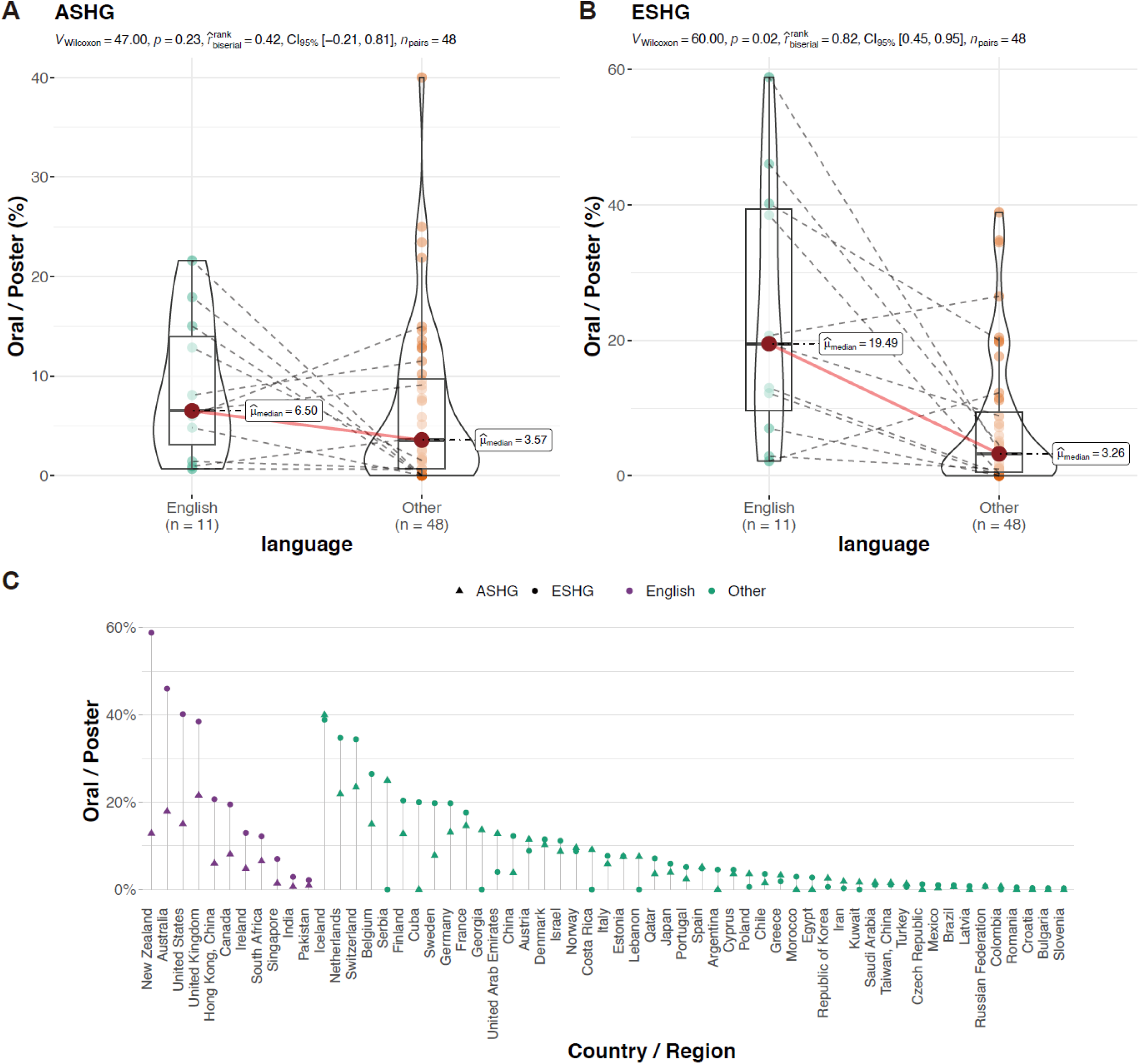
Oral-to-poster ratios by official language status at ASHG and ESHG. Figure A and B show the boxplots comparing countries/regions grouped by English language status, with differences assessed using the Wilcoxon test. Figure C shows oral-to-poster ratios by country/region, also grouped by English language status. Only countries (or regions) with >5 abstracts and non-zero ratios in both conferences are included. Point shape denotes the conference; color indicates whether English is an official language. The y-axis represents the oral-to-poster ratio. We also considered whether English is an official language as a factor influencing the oral-to-poster ratio. After grouping countries based on their English language status, **we found that those where English is an official language had higher oral-to-poster ratios compared to other countries.**

**Supplementary Figure 6.**
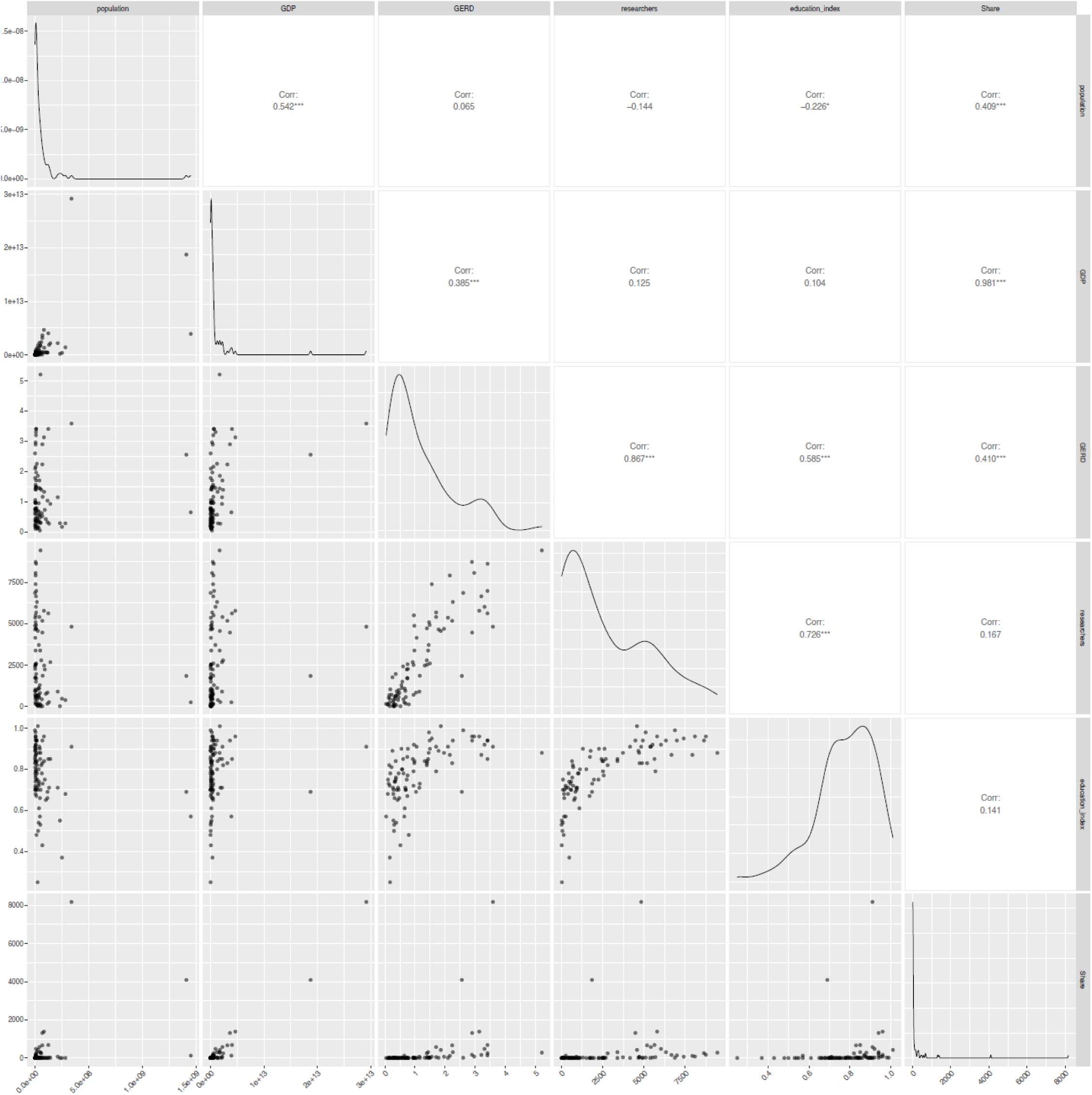
Pairwise plots of six original predictor variables before transformation. This figure displays the pairwise relationships among six original predictor variables: population, GDP, GERD (% of GDP), number of researchers, education index, and Nature Index Share in biological science. The lower triangle shows scatterplots, the upper triangle presents Pearson correlation and significance, and the diagonal shows the distribution of each variable using density curves. **GDP**: gross domestic product (in US dollars). **GERD**: gross domestic expenditure on research and development (in US dollars). **These plots help us visualize raw variable distributions and inter-variable relationships before any transformation.**

**Supplementary Figure 7.**
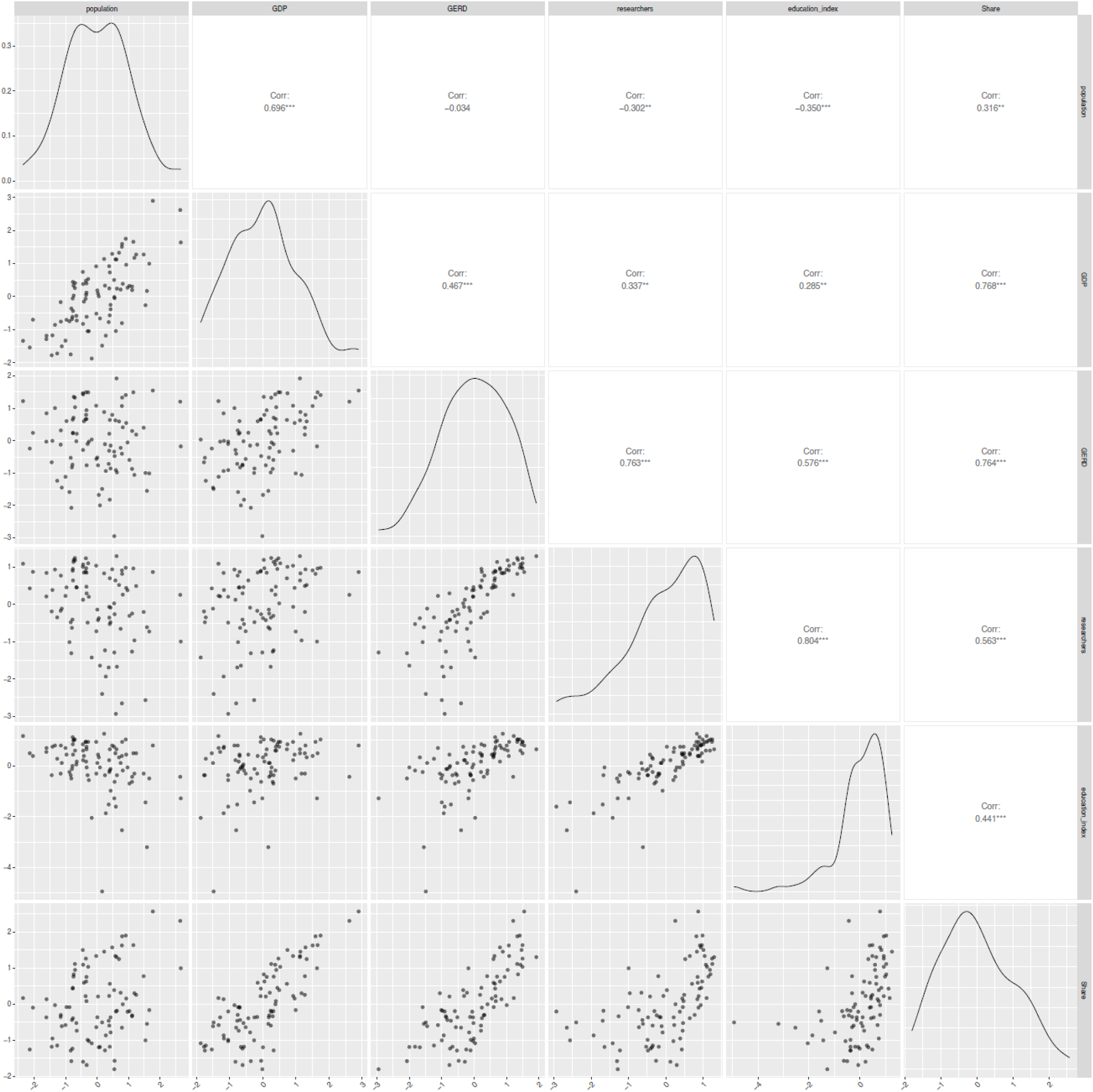
Pairwise relationships of six predictor variables after log_e_ transformation and standardization. This figure shows the relationships among the same six predictor variables after applying a loge transformation followed by z-score standardization. Scatterplots (lower triangle), Pearson correlations (upper triangle), and density plots (diagonal) allow comparison of variable behavior after transformation. **GDP**: gross domestic product (in US dollars). **GERD**: gross domestic expenditure on research and development (in US dollars). **This step helps normalize variable distributions, reduce skewness, and enhance the interpretability and comparability of regression coefficients in downstream analyses.**

